# Strain-stream model of epidemic spread in application to COVID-19

**DOI:** 10.1101/2022.03.26.22272973

**Authors:** S.A. Trigger, A.M. Ignatov

**Affiliations:** Joint Institute for High Temperatures, Russian Academy of Sciences, 13/19, Izhorskaia Str., Moscow 125412, Russia; Institut für Physik, Humboldt-Universität zu Berlin, D-12489 Berlin, Germany; Prokhorov General Physics Institute of the Russian Academy of Sciences, 38 Vavilova St., Moscow, 119991 Russia

## Abstract

The recently developed model of the epidemic spread of two virus stains in a closed population is generalized for situation typical for the couple of strains delta and omicron, when there is high probability for omicron infection enough soon after recovering from delta infection. This model can be considered as some kind of weave of SIR and SIS models for the case of competition of two strains of the same virus having different contagiousness in a population.

**PACS number(s):** 02.50.-r, 05.60.-k, 82.39.-k, 87.19.Xx

## I. INTRODUCTION

Existing models for the spread of infection describe the free-running development of an epidemic and describe all its stages. There are two basic models for such description: the susceptible-infected-susceptible (SIS) models and susceptible–infectious–removed, susceptible–exposed–infectious–removed (SIR, SEIR) models. The SIS model goes back to the pioneering work of Kermack and McKendrick [1] and uses the assumption that the recovered people can immediately get infection again. Existing SIR models assume that the recovered people save strong immunity during epidemic (see, e.g. [2]). There are many variants of those models [3–6] (see also references therein). Balance between the susceptible and infected members of population under the various conditions of infection transfer, are the subject of research in [7–9].

Recently the delayed time-discrete epidemic model (DTDEM) which takes into account typical long duration of the COVID-19 disease has been developed [10]. In [11, 12] this specific delay has been presented in differential form. The delay discussed in [10–12] assumes that a patient is immune, and in this respect fits the SIR model, not the SIS. The considered delay models does not imply the allocation of a separate category of hidden virus carriers (see, e.g., the SEIR models in [13, 14]). Latent carriers of the virus can infect others without delay and are similar to the infected ones. Currently, the simplest SIR and SEIR baseline models are being developed taking into account the vaccination process [15–17].

In the recent paper [18] the SIR-type model was developed for the case of coexistence of two virus strains spreading in the same population. At the same time, due to different contagiousness, a process of replacement of a less contagious virus by a more contagious one takes place, which was quantitatively described in the work. This model assumed that after the end of the disease with any of these strains, the recovered person remains immune for quite a long time to the both types of strains of the SARS-CoV-2 virus. Such long time immunity is typical for SIR models. This property was named “virus orthogonality” in [18] or in application to strains below is named “strain orthogonality”. At the same time, the characteristic details of such a process were revealed, such as the necessary conditions for the emergence of a maximum in the curves describing the current number of virus carriers, a decrease in the peak incidence of a less contagious strain when a more contagious strain appears, a faster depletion of the part of population that has not affected by any of the strains. This means a more rapid course of the epidemic when a more contagious strain appears (if a third, even more contagious strain, does not arise) and increase in the required level of recovered patients to achieve collective immunity (if it turns out to be possible) when a less contagious strain of the virus is replaced by a more contagious one, etc.

The model considered in [18] makes it possible, in the presence of a minimum number of parameters, to quantitatively describe various specific situations of coexistence and struggle of two strains for dominance in a population of living organisms. The specific examples considered in this work were based on the choice of initial conditions that correspond to the emergence of a second strain of high contagiousness (for example, omicron) against the background of an already developed epidemic with the dominance of the delta strain. To describe such a situation, it suffices to take into account the initial conditions, bringing them into line with the actual level of delta disease in a certain population, to the time the strain appeared in South Africa, which was subsequently named omicron by WHO.

At the same time, if we are interested in the rivalry of two strains (for specificity, below we designate delta - 1 and omicron - 2) in any country, region, city or locality, we naturally must use the available statistical data on the incidence, caused by the 1 mutant when cases of the disease caused by the 2 strain appear. For different countries, the corresponding data are quite fully reflected in [19]. City data are presented on the websites of the respective countries (for example, the Robert Koch Institute in Germany, Stopcoronavirus in Russia, the Johns Hopkins Institute in the USA, etc.).

## II. EQUATIONS FOR THE CASE OF “STRAIN NON-ORTHOGONALITY”

In this paper, the basic equations [18] are generalized to the case of “strain non-orthogonality”. These generalization we named “strain-stream” equations. This generalization reflects the observable property to be infected with a high probability by the strain 2 of the COVID-19 disease for those who have already been ill and recovered from infection caused by the strain 1. This means that immunity to strain 2 is not developed (or is only partially developed) after disease caused by strain 1. Obviously, for strains that cause COVID-19 (as well as for influenza viruses), there is only limited period of immunity, however much longer than the average disease duration. In fact, the property of “strain non-orthogonality” means that infection caused by strain 2 (omicron) can appear with some probability even immediately after recovering from the disease caused by strain 1 (e.g., delta). According to our knowledge, the disease COVID-19 caused by two strains which simultaneously coexist in one sick person was not observed (in contrast with the rare cases of COVID-19 and flu). At the same time, according to the existing statistical data after infection by strain 2 infection 1 was not observed. The additional reason for this is a fast disappearance of the less contagious strain, as we demonstrate below.

As in [18] we denote *S* the number of never infected people in a closed population *N, I*_1_ and *I*_2_ are the number of strain carriers of type 1 and 2. Then, equations of the model of “strain non-orthogonality”, which takes into account that after disease caused by strain 1 one can be immediately infected by strain 2 (but not vice versa) are

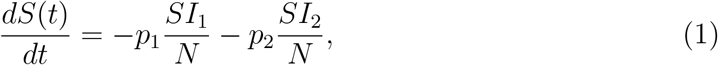

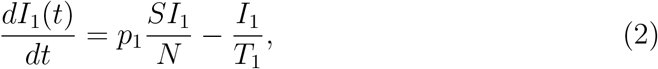

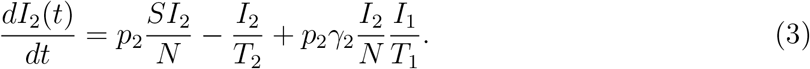

The values *T*_1_ and *T*_2_ are the average durations of the diseases caused by strains 1 an 2. Parameters *p*_1_ and *p*_2_ are the characteristics of the contagiousness for two strains, which are determined as the product of the quantity of dangerous contacts *n*_*c*_ of the infected people per day and the average susceptibility *k* of the healthy person on dangerous distance [10, 11]. The new term in Eq. (3) describes the infection process by strain 2 of the people recovered after the disease caused by strain 1.

The coefficient 0 ≤ *γ*_2_ *<* 1, hereinafter referred to as the Viral Link Attenuation Factor (VLAF), describes a certain decrease in the probability of getting 2 after being infected with 1 (partial increase in immunity) compared to the probability of getting 2 without having been ill before 1 (i.e., directly from the group *u*). This is due to the production of antibodies after the disease caused by the 1 strain, which perform some protective function against the 2 strain (or after vaccination). The structure of the last term in (3) is obvious if we take into account that the proportion of strains 1, 2 recovered from diseases is equal to *R*_1,2_ = *−y*_1,2_*/T*_1,2_ respectively.

In the general case, passing to symmetric equations, we can consider the situation when after the disease 2 it is possible to get sick 1 with a certain probability *γ*_1_, but this mathematical generalization is not considered in this article as unrealizable for omicron and delta strains.

Equations (1)-(3) describing the epidemic spread for the case of two “non-orthogonal” strains in the closed population *N* can be rewritten in the form using the variables *I*_1_(*t*)*/N* = *y*_1_(*t*), *I*_2_(*t*)*/N* = *y*_2_(*t*), *S/N* = *u*(*t*)

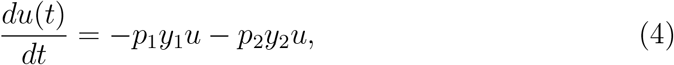

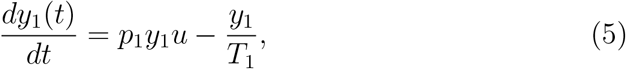

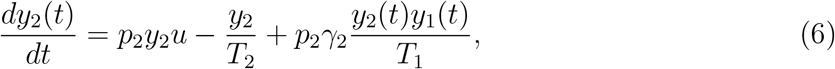

Here we use the same notations as in [18]. The value *u*(*t*) ≡ 1 − *z*(*t*) is the fraction of the population that is not affected by virus at all, *z*(*t*) = *N*_*tot*_(*t*)*/N* corresponds to the fraction of full population *N* that are affected (ill and recovered *N*_*tot*_(*t*) ≡ *N*_1_(*t*) + *N*_2_(*t*)) by the strain 1 (*N*_1_) or the strain 2 (*N*_2_) to the time *t*. The values *y*_1_(*t*) = *I*_1_(*t*)*/N* and *y*_2_(*t*) = *I*_2_(*t*)*/N* are the current fractions of population actively infected (viruses carriers) by strains 1, 2 respectively in a moment *t*.

The two-strain propagation model developed in [18] is the limiting case of the considered more general model (1)-(3) for *γ*_2_ = 0. We also use the *x*_*i*_ values for the proportion of those affected (recovered and sick) by the strain *i* = 1, 2 to the moment of time *t*

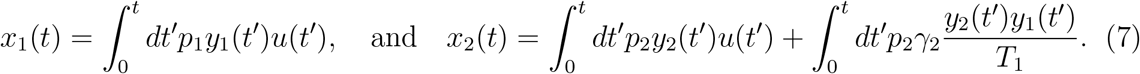

## III. NUMERICAL SOLUTION FOR VARIOUS IMMUNITY PARAMETER VLAF

An analysis of the stability of the stationary solution, carried out in [18], showed that the necessary condition for the development of an epidemic process at *γ*_2_ = 0 is the condition *p*_*i*_*T*_*i*_*u*_0_ > 1. This condition remains valid for equations (4)-(6).

As was revealed in [18] for *γ*_2_ = 0, using the example of specific initial conditions and parameters *p*_*i*_ and *T*_*i*_, the coexistence of two viruses of different contagiousness leads over time to the replacement of the less contagious strain by a more contagious one, even if the share of the latter at the beginning of the process was significantly smaller than less contagious. The results of calculations for specific parameters that correspond to the simultaneous emergence of an epidemic with two strains of different contagiousness are shown in Fig. 1 (epidemic process in the presence of one strain 1, when *p*_2_ = 0) and Fig. 2 (comparison of the dynamics of the epidemic in the presence of both strains for the model under consideration with *γ*_2_ = 0.3.

**Figure 1:**
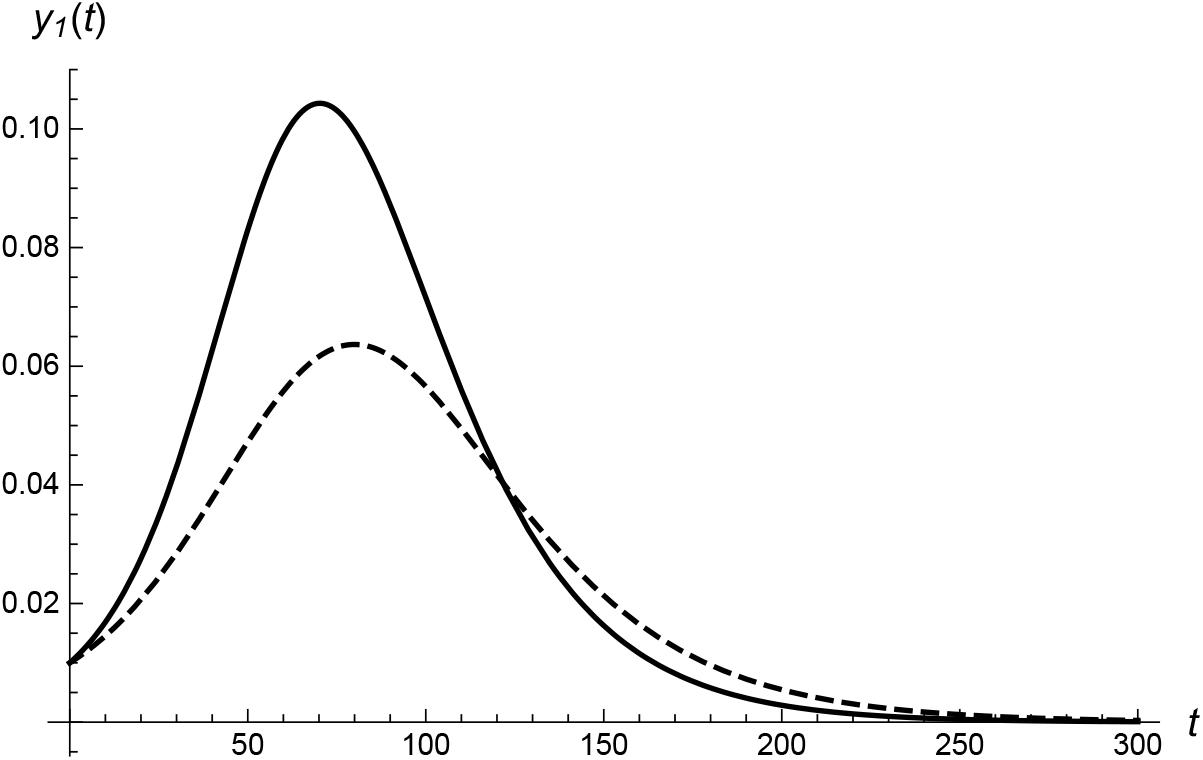
Function *y*_1_(*t*) of strain 1 carriers, when the second strain is absent (*p*_2_ = 0) for *u*(0) = 0.8 (solid) and for *u*(0) = 0.7 (dashed). The parameters are *p*_1_ = 0.15, *y*_1_(0) = 0.01, the average duration of the virus carrier *T*_1_ = 15 days.

**Figure 2:**
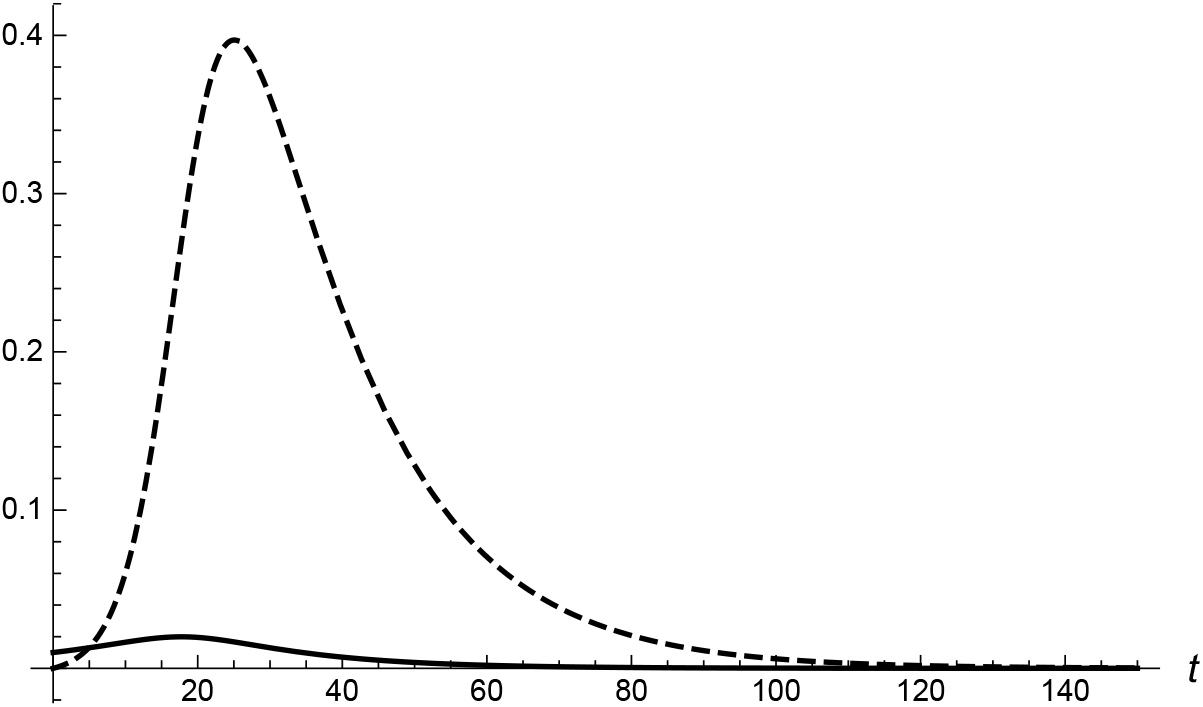
Functions *y*_1_(*t*) (solid) and *y*_2_(*t*) (dashed) of the virus carriers for the case when both strains exist. The parameters are *p*_1_ = 0.15, *p*_2_ = 0.4, *y*_1_(0) = 0.01, *y*_2_(0) = 10^*−*7^, *u*(0) = 0.8, the average duration of the virus carrier *T*_1_ = *T*_2_ = 15 days, *γ*_2_ = 0.3.

Thus Fig. 1 and Fig. 2 serve to demonstrate the process of mutual influence of strains during the development of an epidemic.

As is easy to see that strain 1 is effectively suppressed by strain 2 since the value of maximum for the solid curve in Fig. 2 is approximately five times lower than in Fig. 1 for *u*(0) = 0.8. Comparison of these figures shows that duration of strain 1 circulation is also effectively suppressed (≃ 4 times shorter for the used parameters) due to the appearance of strain 2. Comparison of Figs. 1 and 2 shows that circulation of strain 2 is essentially shorter than in the case of it absence. It is easy to see that for arbitrary parameters the maximum for strain 1 in Fig. 2 is shifted to earlier time in comparison with Fig. 1. This property, mentioned in [18], is valid also for the strain-stream model under consideration.

In this paper, we are interested in the impact of a possible infection with virus 2 after recovery from an infection caused by virus 1. This situation corresponds to the epidemic process observed with the appearance of the omicron strain. An important difference from the specific examples considered in [18] is the appearance of strain 2 under conditions of a developed epidemic of strain 1, which is characterized by rather large initial values of *u*(0) and *y*_1_(0).

The results of the numerical solution of equations (5-7) for the initial conditions simulating the situation of the appearance of omicron in already developed epidemic of the delta strain are shown in Fig. 3 - Fig. 5.

**Figure 3:**
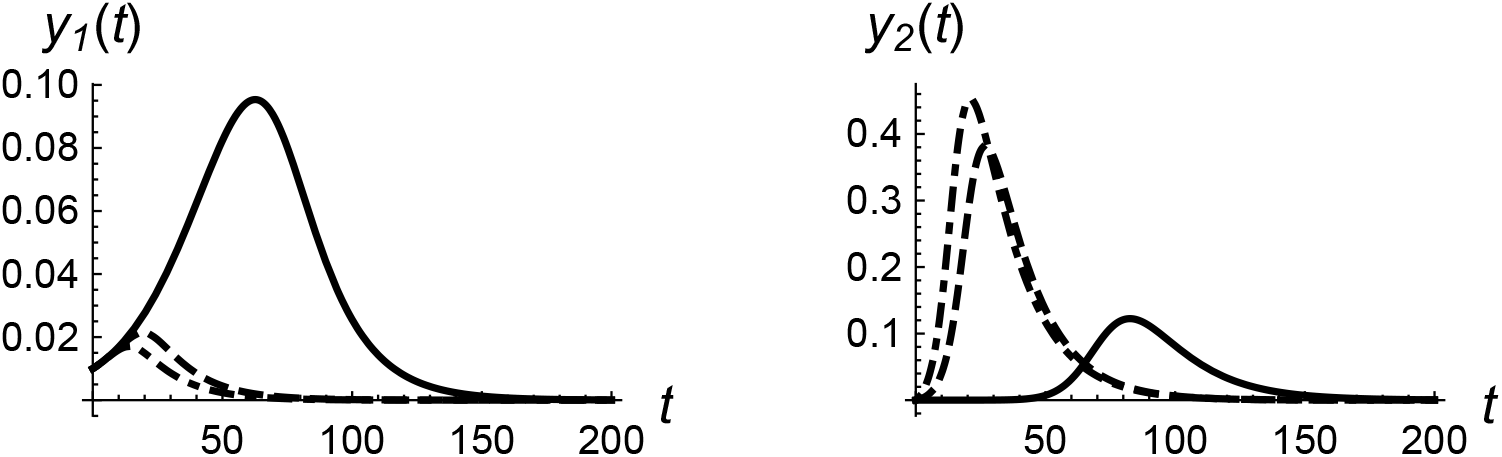
Comparison of the function *y*_1_(*t*) (left), and virus *y*_2_(*t*) (right) for different values *γ*_2_ = 0 (solid), *γ*_2_ = 0.2 (dashed) and *γ*_2_ = 0.8 (dash-dotted) of virus carriers for the case when both virus strains exists. The parameters are *p*_1_ = 0.15, *p*_2_ = 0.4, *y*_1_(0) = 0.01, *y*_2_(0) = 10^*−*7^, *u*(0) = 0.8, the average duration of the virus carrier *T*_1_ = *T*_2_ = 15 days.

The proportions of *y*_1_(*t*) and *y*_2_(*t*) infected with strains 1 and 2 are shown in Fig. 3 for different parameters *γ*_2_, left and right, respectively. As in Fig. 1 and Fig. 2, the initial condition for the proportion of the population that did not encounter either of the two considered strains was chosen at the level of *u*(0) = 0.8, which significantly exceeds the official statistics for, e.g., Germany at the time the omicron strain appeared in the country. By such an overestimation, we take into account a significant number of unreported cases of diseases with the delta strain at the time of the appearance of the omicron strain. The same qualitative picture is observable also in other countries. The initial proportion of those infected with strain 1 is chosen to be very high *y*_1_(0) = 0.01, which also corresponds to the presence of a significant number of hidden virus carriers that can actively infect others. Note, that the purpose of this work is to identify the general patterns of the development of the epidemic in the presence of two strains, and not a calculation based on a detailed analysis of the changing situation from day to day and incomplete statistical data.

As follows from Fig. 3, the impact of the appearance of strain 2 capable of infecting those who have been ill with strain 1 depends significantly on the value of VLAF *γ*_2_. The more 0 ≡ *γ*_2_ ≡ 1, the faster the process of infection with strain 1 is suppressed, i.e. it is forced out faster than in the original model with *γ*_2_ = 0 [18] (see also Fig. 1). At the same time, as *γ*_2_ grows, the current proportion of strain 2 carriers grows, exceeding by a factor of 4.5 at the maximum proportion of strain 1 carriers under the chosen parameters.

The effect of a non-zero value *γ*_2_ on the fraction *u*(*t*) of non-affected by strains at all is shown on Fig. 4. Possibility to become infected with strain 2 soon after disease caused by strain 1 is high. There is a much faster and complete depletion of the share of non-affected. This means that with a certain parameter *γ*_2_, herd immunity becomes practically unattainable and almost everyone must get sick due to strain 2. It is of interest to determine values *γ*_2_ for which the stationary value of the proportion of the population not affected by any of the viruses is reached. It can be considered as a numerical characteristic of herd immunity. The calculation carried out up to 1000 days (not shown in Fig. 4) showed that with the selected parameters, the solid curve corresponding to the absence of strain 2 tends to *u*(*t* = 1000) = 0.205, the dash-dotted curve (corresponding to the case *γ*_2_ = 0 [18] of full lengthy in time immunity after each of the diseases caused by the strain 1 or 2) tends to 0.033 and the dotted curve corresponding to the case under consideration Eqs. (4)-(6) for *γ*_2_ = 0.2 tends to *u*(1000) = 0.005. In the latter case the level of collective immunity is only 0.5 percent of the population.

**Figure 4:**
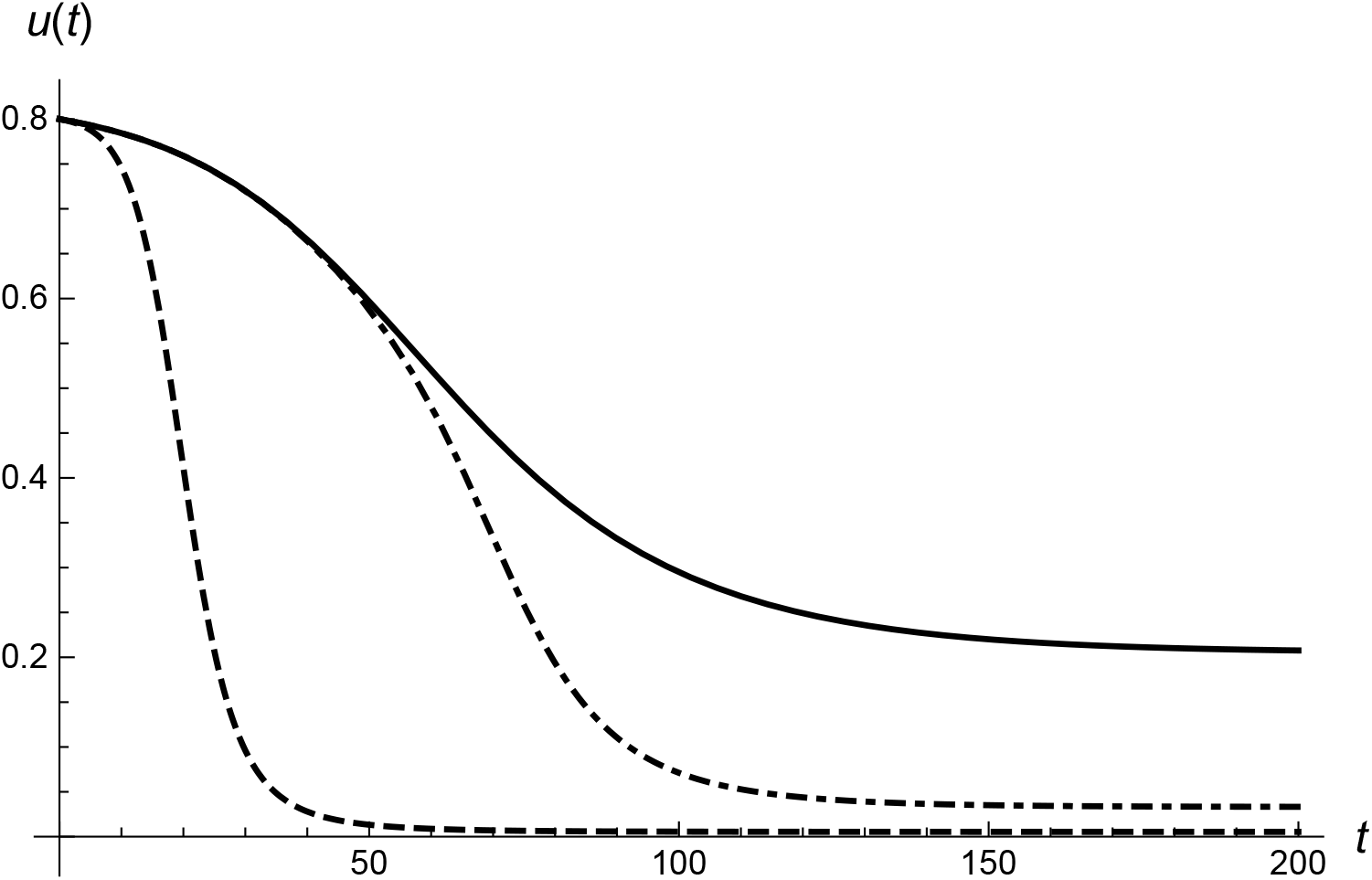
Function *u*(*t*) for the various cases: the second strain is absent (*p*_2_ = 0), the parameters are *p*_1_ = 0.15, *y*_1_(0) = 0.01, the average duration of the virus carrier *T*_1_ = 15 days for *u*(0) = 0.8 (solid); both strains coexist, the recovered are immune (the case considered in [18]), the parameters are *p*_1_ = 0.15, *p*_2_ = 0.4, *y*_1_(0) = 0.01, *y*_2_(0) = 10^*−*7^, *T*_1_ = *T*_2_ = 15 days for *u*(0) = 0.8 (dash-dotted), *γ*_2_ = 0; both strains coexist, the parameters are *p*_1_ = 0.15, *p*_2_ = 0.4, *y*_1_(0) = 0.01, *y*_2_(0) = 10^*−*7^, *T*_1_ = *T*_2_ = 15 days for *u*(0) = 0.8, *γ*_2_ = 0.2 (dashed).

Fig. 5 shows the curves for the total part of people (sick plus recovered, or affected) *x*_1_(*t*) with strain 1 (left) and strain 2 (right), calculated according Eq. (7). All three curves for the function *x*_1_(*t*) (left) and for the function *x*_2_(*t*) (right) in Fig. 5 correspond to the circulation of two strains, but for different values *γ*_2_. The parameters in Fig. 5 correspond to those selected in Fig. 3. As follows from Fig. 5 the function *x*_1_(*t*) decreases as *γ*_2_ increases, while the function *x*_2_(*t*) grows. This behavior corresponds to the general pattern of replacement of a less contagious virus by a more contagious one, with the greater efficiency, the greater the VLAF value.

**Figure 5:**
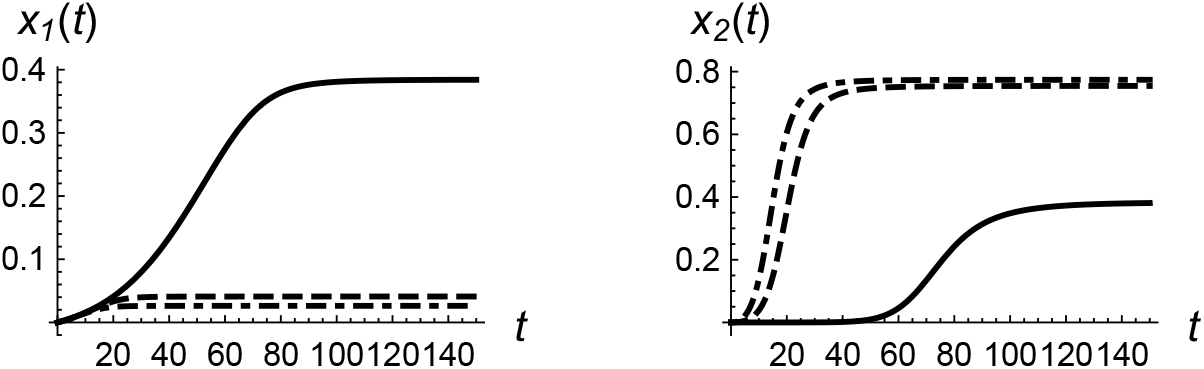
Function *x*_1_(*t*) (left) of the people affected by the strain 1 and *x*_2_(*t*) (right) of the people affected by the strain 2 for different values *γ*_2_ = 0 (solid), *γ*_2_ = 0.2 (dashed) and *γ*_2_ = 0.8 (dash-dotted) of virus carriers for the case when both virus strains exists. The parameters are *p*_1_ = 0.15, *p*_2_ = 0.4, *y*_1_(0) = 0.01, *y*_2_(0) = 10^*−*7^, *u*(0) = 0.8, the average duration of the virus carrier *T*_1_ = *T*_2_ = 15 days.

The formulated equations and the model under consideration can be easily extended to take into account vaccination and different quarantine measures, accounting the government restrictions, vaccination process etc. In fact, after recovering from strain 1, a person may not immediately become infected with strain 2, and taking into account this factor associated with time shifts is beyond the scope of this work. Also the cases of death, re-infection with the same strain long time after recovery, limited time for vaccination efficiency and other known factors can be included in a more elaborated models. Above we restricted our consideration by the case of the free running epidemic under two “non-orthogonal” strains of a same virus. This assumption can be considered as realistic for very fast developing epidemic caused by, e.g., the omicron strain (or another highly contagious virus strain) appeared in a population affected earlier by a less contagious virus strain. However, the considered model clarifies the main specific features of competition of two “non-orthogonal” viruses in population.

## IV. CONCLUSIONS

The principal picture of the replacement of one strain by another has already been revealed in the recently considered new mathematical model [18], where the basic equations were proposed that describe the replacement of a less contagious virus by a more contagious one. Further development of the theory is connected with taking into account the incomplete “orthogonality” of the strains under consideration. This is manifested in the fact that with a significant mutation of the virus, leading to a different molecular structure, a different virulence, and a different clinical picture of the disease, both strains, spreading in the population, are mutually more dependent. Immunity to one of them (for example, due to a previous disease), generally speaking, does not means the presence of immunity in relation to another. So, for example, omicron can infect those who have recovered from the delta strain, but not vice versa.

Thus, the situation cannot be described in the framework of SIR and similar models, where all recovered patients have a long immunity, nor within the SIS model, where immunity disappears immediately after recovery. This important property is taken into account by transferring to the “strain-stream” equations by modification of Eq. (3) and respectively (6). The additional term includes the new VLAF parameter *γ*_2_ ≤ 1, due to the development of partial immunity to strain 2 as a result of the disease caused by strain 1, or to the effective vaccination against strain 1, giving partial protection also against strain 2.

## Data Availability

All data produced in the present study are available upon reasonable request to the authors

https://doi.org/10.1101/2022.01.11.22269046

## Acknowledgment

S.T. is thankful to infectious disease physician Dr. M. Karavaeva, Prof. Dr. F. Onufrieva and colleagues from the clinic Charité (Berlin) for many useful discussions.

